# Dynamical repertoire of brain networks in mindfulness cognitive behavioural therapy during rumination: A randomized controlled trial

**DOI:** 10.1101/2025.02.04.25321569

**Authors:** Anne Maj van der Velden, Jakub Vohryzek, Willem Kuyken, Jesus Montero-Marin, Guusje Collin, Morten L. Kringelbach, Henricus G Ruhe

## Abstract

**BACKGROUND:** Depression is a prevalent and debilitating affective disorder characterised by the dominance and persistence of depressive rumination. Mindfulness-based cognitive therapy (MBCT) is an effective treatment for recurrent depression developed specifically to target rumination and recurrence risk by training metacognitive awareness and adaptive attention, emotion and self-regulation skills. Yet, the underlying mechanisms by which mindfulness training impacts maladaptive depressive rumination is not well understood, and a deeper understanding of its effects on the complex brain dynamics during depressive rumination is needed.

**METHOD:** In a randomised controlled functional magnetic resonance imaging (fMRI) study (N = 80), we used LEiDA (Leading Eigenvector Dynamics Analysis) to determine the key substates during resting state fMRI of an experimentally induced rumination state before and after treatment with MBCT (N = 27) for recurrent depression in addition to treatment as usual (TAU) or TAU alone (N = 21). We determined the probability of occurrence (fractional occupancy) and the duration (lifetime) of underlying substates (phase-locking patterns) before and after treatment for both groups.

**RESULTS:** We found that MBCT training compared with TAU altered the fractional occupancy of a ‘Salience-somatomotor’ substate during the depressive rumination induction. These dynamic network changes in turn were associated with reduced depressive symptoms after treatment and at three months follow up.

**CONCLUSION:** In a depressive ruminative state, changes in the dynamics of the somatosensory-salience network following mindfulness training was associated with improved clinical outcomes which provides insight into candidate brain mechanisms or markers of treatment response.

**Trial registration:** ClinicalTrials.gov <u>NCT03353493</u>.

## Introduction

Depression is a leading cause of disability worldwide (1). One of the most debilitating aspects of depression is the dominance and persistence of depressive rumination (2). Depressive rumination is characterised by a persistent, rigid and self-related focus on depressed mood and its causes and consequences (3). Rumination has been found to predict increased risk of recurrence in remitted individuals, severity of depressive episodes, and risk of developing treatment-resistant and chronic depression trajectories (2,4,5).

Mindfulness-based cognitive therapy (MBCT) is an effective treatment for recurrent depression and is recommended as a preventive treatment in a number of national health guidelines (6–8). As with other effective treatments for recurrent depression, adding MBCT to Treatment as Usual (TAU) can approximately half the risk of relapse, but many individuals will remain vulnerable to experience residual depressive symptoms, relapse, or recurrence of a full depressive episode. To optimize outcomes, we need to develop a greater understanding of why, when and for whom MBCT+TAU works (9). MBCT was developed to target depressive rumination and recurrence risk by training meta-cognitive awareness and adaptive attention, emotion and self-regulation skills (10). More specifically, participants are taught to recognize, decenter and disengage from ruminative negative thoughts, by redirecting attention to the embodied experience of present-moment sensations, and relate to the changes in present-moment experience with a non-judgmental and accepting attitude (11). Yet, the underlying mechanisms by which mindfulness training impacts maladaptive depressive rumination is not well understood, and hence a deeper understanding of how MBCT impacts complex neurocognitive processes during depressive rumination is warranted.

Mindfulness-based interventions and practices have been found to alter brain function in regions and circuits that underlie attention, interoception, emotion regulation, and self-relevant processing (for reviews, see (12–15)). Areas in the salience network and default mode network have especially been implicated in rumination, mindfulness training and depressive symptomatology (2,12,15–17). However, research on the neural correlates of MBCT for recurrent depression and depressive ruminative mind states is still very scarce (18). To our knowledge, in the first neurocognitive study on depressive rumination in relation to MBCT a, we found that MBCT+TAU compared to treatment as usual (TAU) led to decreased salience network connectivity to the lingual gyrus during a ruminative state, and that this change in salience network connectivity mediated improvements in the ability to sustain and control attention to body sensations (18) using static connectivity measures. While these findings are indicative that reduced salience network connectivity might play a role in how likely participants are to get stuck in persistent ruminative processing, static connectivity measures cannot elucidate how stable or variable the connectivity patterns are over time, which we hypothesise to be key to understand and disentangle the complex and subtle neural dynamics during depressive rumination (4).

Progress has been made in the ability to characterize functional connectivity at the whole-brain scale from a dynamic, rather than static, perspective (19). A dynamic approach (20–24) can elucidate the flexibility or rigidity of specific brain substates during a ruminative mind state (25). Hence, functional dynamic connectivity methods may yield valuable insight into how neural dynamics during a ruminative state may change after a mindfulness-based intervention, in a way that cannot be captured by traditional static functional connectivity measures looking at an average connectivity (4).

Thus, we aim to investigate how MBCTimpacts neurocognitive dynamics during depressive rumination by applying dynamic functional connectivity in a secondary analysis of a randomised controlled fMRI trial with MBCT+TAU vs. TAU for recurrent depression compared to treatment as usual. Specifically, we aim to investigate how mindfulness training impacts the probability of occurrence and duration of the key brain substates present in a depressive ruminative mind state, and whether changes in these neural dynamics are associated with clinical outcomes.

## Methods and Materials

### Study design and participants

The trial has been fully described elsewhere (18). In brief, we conducted a randomised controlled trial examining change in neurocognitive functioning between individuals participating in Mindfulness-Based Cognitive Therapy in addition to treatment as usual (MBCT+TAU) or TAU only. The study was registered at ClinicalTrials.gov Identifier: NCT03353493, and was approved by the Regional Ethics Council of Central Jutland, Denmark.

Eighty adult participants with a diagnosis of recurrent major depressive disorder with or without a current episode were recruited and gave written informed consent. An independent researcher randomly allocated (5:3 ratio) participants to receive either an 8-week MBCT class + TAU treatment or adhere to TAU treatment alone using a computerised system. Randomization was stratified according to antidepressant usage and participants’ symptomatic status. The researchers conducting MRI scans were masked to treatment allocation, and questionnaires were administered online.

### Intervention

MBCT+TAU is an 8-week manualized group-based intervention, which combines psychoeducational elements from cognitive behavioural therapy for depression with a systematic training in mindfulness meditation. The programme consists of weekly classes of 120 mins with daily homework in between. The programme was taught in university settings by highly experienced therapists fulfilling internationally recognized ‘good practice’ guidelines for teachers, trainers and supervisors of mindfulness courses (26). TAU was restricted to no psychotherapeutic intervention and either a stable dose of antidepressant medication or no medication. Participants in the TAU group were offered MBCT after they completed all the study assessments.

### Measures and procedures

Participants were assessed at baseline (before randomization), after the end of the 8-week MBCT program or 8 weeks after baseline for TAU-only participants, and after 3 months of follow-up. Each assessment consisted of questionnaires and MRI scanning sessions.

### Psychological processes and clinical measures

We assessed depressive symptoms using the Quick Inventory of Depressive Symptomatology (QIDS_SR (27)); perceived stress using the Perceived Stress Scale (PSS (10)); interoceptive awareness using the subscales of noticing (N0), emotional awareness (EA), body listening (BL), attention regulation (AR), trusting (TR) and not-distracting (ND) of the Multidimensional Assessment of Interoceptive Awareness (MAIA) (28); decentering using the Experiences Questionnaire (EQ) – decentering factor (29); mindfulness skills using the Five Factor Mindfulness Questionnaire short form (FFMQ-SF (30)), and trait rumination using the Rumination Response Scale (RRS (4)). All questionnaires were administered before and after treatment or at 8 weeks in the TAU participants.

### fMRI paradigm and rumination induction

The fMRI paradigm on depressive rumination, was part of a larger paradigm including an initial structural scan followed by four separate functional connectivity scans (5 minutes each) in the consecutive order of resting state, an instructed mindfulness state, resting state, and an instructed rumination state. In this paper we focus on the changes in the rumination induction state, as our aim is to explore whether, and if so how, MBCT+TAU impacts neural dynamics of probability of occurrence and duration of key substates during a ruminative mind state.

In the rumination induction state, participants were guided through a rumination induction adapted from (19) in which participants first rehearsed a sad autobiographical memory and subsequently were instructed to stay with their sad mood and reflect on self-related causes and consequences of their low mood. Participants were free to select their own sad autobiographical memory. Before undergoing scanning at each timepoint, the research team took care to instruct participants about the nature of the task and highlight its voluntary nature for ethical reasons. In addition, there was a brief follow-up talk with a clinically trained member of the research team; the purpose of which was to make sure participants were okay after the experiment. Participants were scanned at baseline and within a month after treatment.

### MRI Acquisition and fMRI Preprocessing

Structural and functional images of the brain were acquired on a 3T Siemens Magnetom Skyra 3T scanner with a 32-channel head coil (using software version Scout; Siemens Healthineers). For preprocessing, we used FSL tools (version 6.0). Preprocessing steps followed standard procedures including i) registering the functional to the structural image, ii) registering the structural image to standard space, iii) motion correction, iv) spatial smoothing, and v) high-pass filtering (100-second cutoff). For full details for the preprocessing steps, see the Extended Methods and Materials in the Supplement).

### Leading Eigenvector Dynamics Analysis

In this study, we applied Leading Eigenvector Dynamics Analysis (LEiDA) in order to examine dynamic network changes in a ruminative brain state in patients with recurrent depression undergoing MBCT treatment. More specifically, we aimed to elucidate the properties of the spatial neural substates, namely Fractional Occupancy (the percentage of frames assigned to a substate for the whole of the rumination scan) and Lifetimes duration (the mean duration of temporally continuous runs of substate occupancy). For each participant and condition separately (pre- and post-intervention scans in the ruminative state), we transformed a parcellated 90 regions (AAL-based atlas) fMRI signal into an analytical signal, which allowed us to compute the phase relationship between each pair of regions at every recorded time point of the pre and post conditions (See Figure 1 for overview of the study design and the analytical approach). We subsequently computed the leading eigenvectors of each phase-locking pattern (a pattern consisting of regions with similar phase) at every recorded timepoint. Then, applying an unsupervised k-means clustering algorithm (a machine learning tool to find structure in the data), we defined the number of clusters based on i) optimal clustering performance as determined by a silhouette score (a control measure quantifying the cohesion of the clusters) and ii) the lowest clustering solution that would result in a Bonferroni corrected statistical significance.

**Figure 1:**
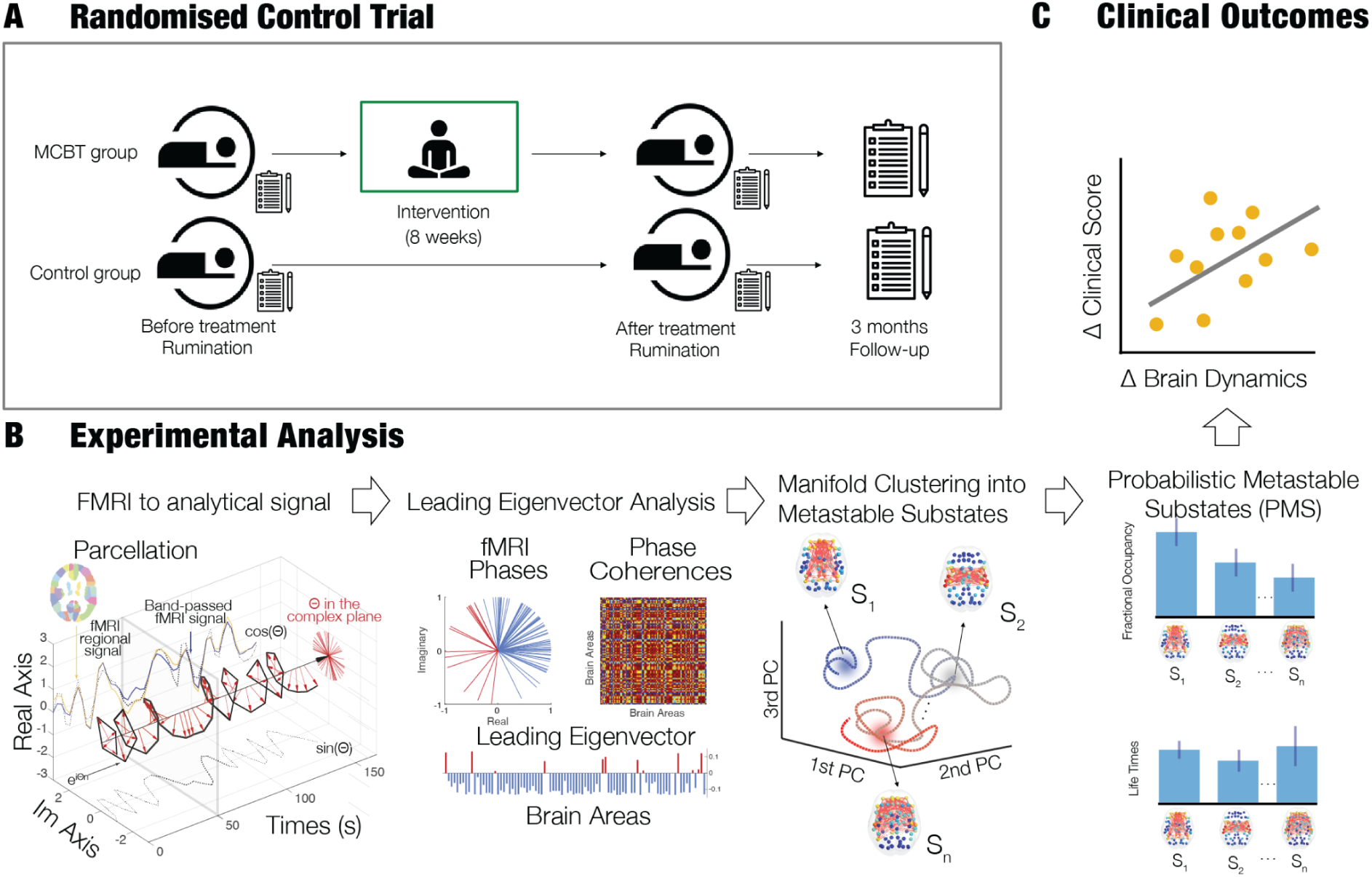
Experimental Design and Leading Eigenvector Decomposition Analysis (LEiDA) pipeline. **A)** Study design including randomization, fMRI measures at baseline and post treatment and clinical data collection at baseline, post-treatment and 3 months follow up. **B)** The Automated Anatomical Labelling (AAL) atlas parcellated fMRI time series are transformed into the analytical signal. Instantaneous phase is extracted for each region at every time point of the recording, instantaneous phase coherence matrix is computed and decomposed in order to obtain the leading eigenvectors for each timepoint of the recording. Unsupervised clustering is performed to obtain the LEiDA substates and then dynamic measures of Fractional Occupancy and Lifetimes Duration can be computed. Figure adapted from (21). **C)** Comparing change in fMRI measures (pre and post treatment) with changes in depressive symptoms (pre and post treatment and 3 months follow up).

To thoroughly investigate different partitions, we gradually varied k from 2 to 20. The number of clusters, k, in the k-means clustering algorithm determines the range of phase-locking (PL) substates in the repertoire. By increasing k, more intricate, infrequent, and asymmetric substates are revealed. This approach allowed us to analyse substates that exhibited significant differences in brain activity between the intervention and control groups. Lastly we computed the dynamic measures (fractional occupancy and lifetimes duration) for each phase-locking substate, comparing change from pre to post treatment in the MBCT+TAU and TAU groups. See extended methods for further details of the experimental analysis (SX)

### Comparisons with psychological processes and clinical outcomes

We compared the statistically significant changes in fractional occupancy and lifetimes duration with significant changes in psychological processes and clinical outcomes, and followed up significant correlations up with regression analysis looking at whether changes in dynamic brain measures were associated with changes in psychological processes and clinical outcomes and whether such a relationship was specific to the MBCT+TAU group compared with TAU, and as such may provide insight into candidate neural mechanisms or markers of treatment response.

## Results

For the study, 80 participants were randomly allocated to receive MBCT+TAU (n = 50) or TAU alone (n = 30) (Figure 1). We had 48 subjects (27 MBCT+TAU; 21 TAU) completing post scanning sessions including the rumination condition and obtaining the full treatment (Figure 1). The baseline characteristics were similar between the two groups (18).

Out of ethical concerns, the rumination condition of the fMRI paradigm was voluntary. The participants not undertaking the rumination condition (n = 20) had similar baseline characteristics except for higher depressive symptoms (18).

We chose the number of substates as K=8 by examining the optimal clustering performance as determined by the silhouette score as well as the lowest clustering solution that resulted in Bonferroni corrected statistical significance for the Fractional Occupancy measure (see supplementary Figures S2 and S3)

### Analysing changes in fractional occupancy and lifetimes duration during rumination

We first analysed the Fractional Occupancy and Lifetime duration for the clustering solution k = 8. For each partition of the 8 substates (horizontal axis). The p-values resulting from between-group comparisons (using the independent samples t-test) of the post-minus pre-treatment Fractional Occupancies and Lifetimes (i.e. change over time) per group are shown in Figures 2, 3 and S4. For the chosen clustering solution of 8 substates, a significant decrease in the Fractional Occupancy of Salience-somatomotor substate (p-val = 0.044) was observed after correcting for multiple comparisons for MBCT+TAU vs. TAU (Figure 2A). This Salience-somatomotor substate consisted of bilateral areas of the salience network (i.e. the insula) and several areas of the somatosensory and motor networks (i.e rolandic operculum, supramarginal gyrus, frontal inferior operculum, temporal superior gyrus, precentral and postcentral gyrus; putamen), but also auditory areas (i.e. Heschl’s gyrus; areas of the temporal pole (Temp pol sup)) and part of the basal ganglia such as the pallidum (Figure 3). This difference in the Fractional Occupancy of this substate was consistent over a range of clustering solutions k=7 to k=20 (except k=11 and k=17), with k=8 and k=9 showing the most robust differences for the Bonferroni corrected p-values.

**Figure 2:**
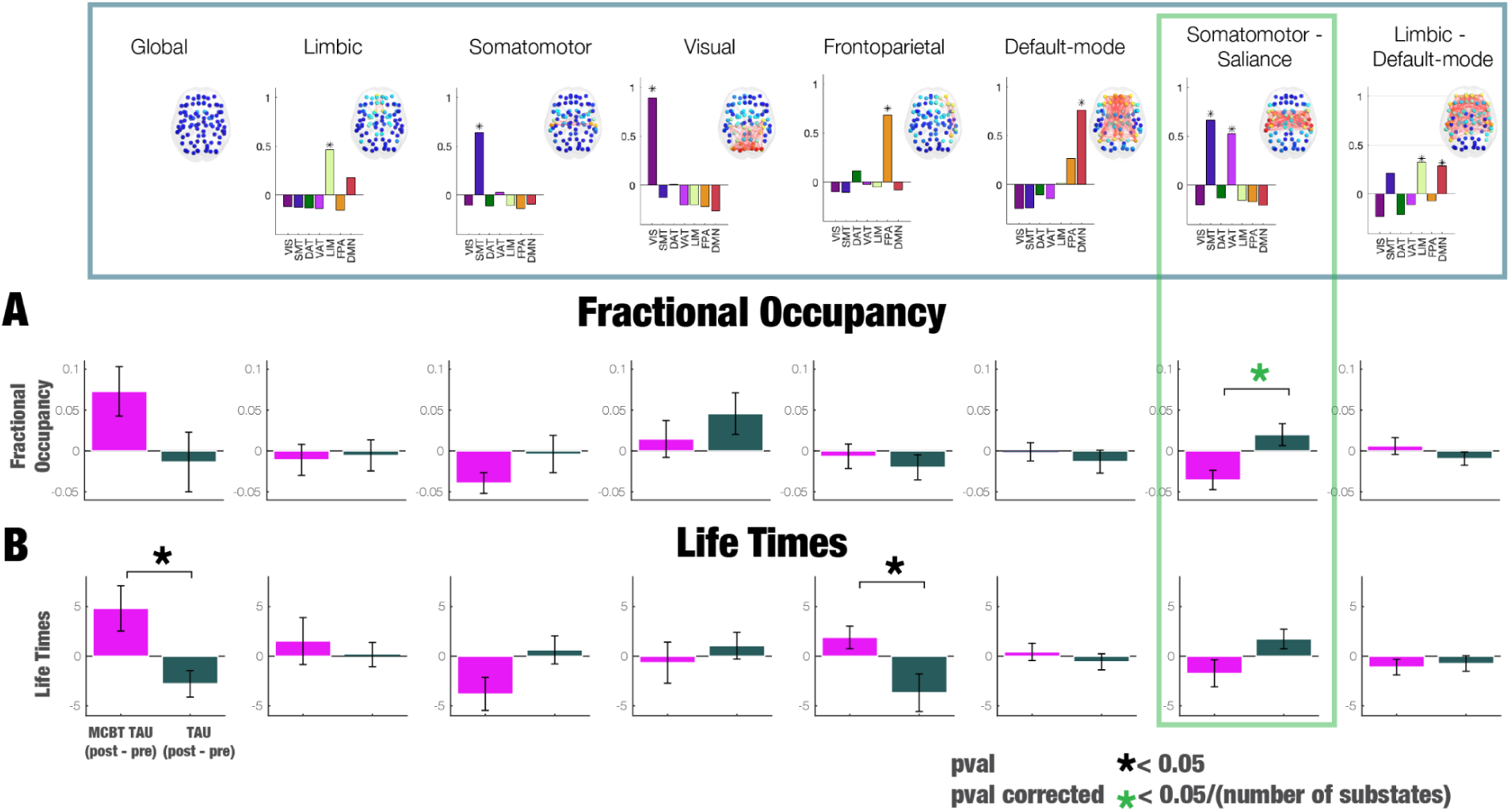
Repertoire of recurrent phase-locking substates in neuroimaging data of MBCT+TAU and TAU groups. **A)** Chosen clustering solution of 8 substates. We found a significant decrease in Fractional Occupancy of a Salience-somatomotor substate (p-val = 0.0055, corrected for multiple comparisons across 8 substates (p-val = 0.044)) comparing MBCT+TAU pre-post vs TAU pre-post. **B)** We found a trend of an increase in Lifetimes duration of the global substate and the frontoparietal substate (p-val = 0.012 and p-val = 0.019 respectively, uncorrected for multiple comparisons) comparing MBCT+TAU pre-post vs TAU pre-post. Assessment of the difference between groups was performed with independent samples T-tests, in this specific clustering solution (k=8) for Fractional Occupancy and Lifetimes Duration. Black “*” signs significant uncorrected p-values (<0.05), green “*” signs significant Bonferoni corrected p-values (<0.05/8).

**Figure 3:**
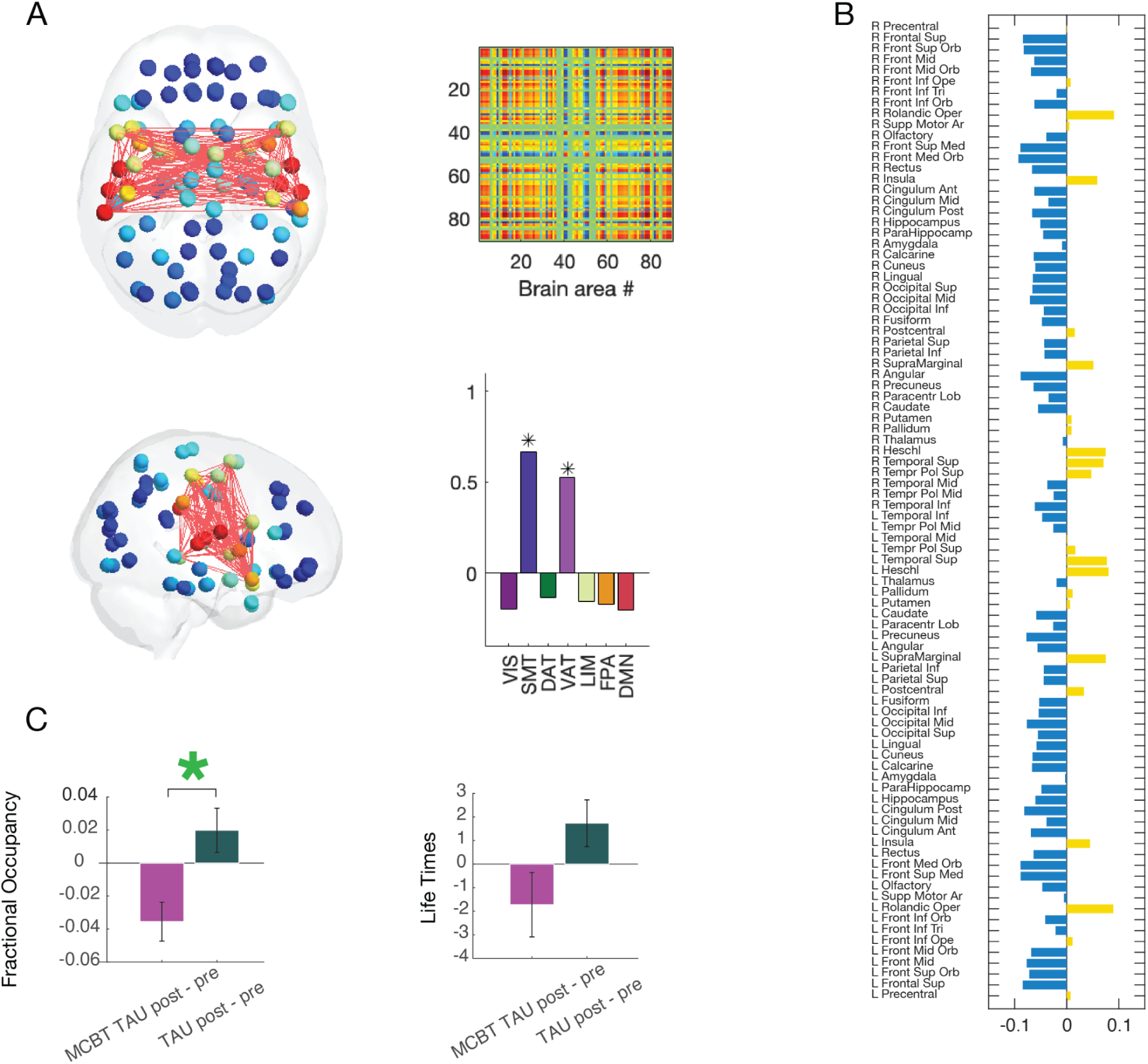
Change in the Fractional Occupancy of the Salience-somatomotor substate. **A)** Cortical renderings of the Salience-somatomotor substrate (7th phase-locking substate in the K=8 clustering solution). The matrix represents coherence between AAL nodes and their rendering on the resting-state networks as defined by Yeo et al. (2011) (31). **B)** Cortical labels for the Salience-somatomotor substate (in yellow the regions associated with this substate). **C)** Decrease in Fractional Occupancy of the Salience-somatomotor substate after MBCT+TAU+TAU versus an increase in the TAU group. No differences between groups were found for the Lifetime duration of this substate.

A trend toward increased Lifetimes duration of the global substate (clustering solution substate 1; p-val = 0.012) and a frontoparietal substate (p-val = 0.019) was observed (significant p<0.05 before corrections for multiple comparisons; Figure 2B).

### Relating change in fractional occupancy during rumination to psychological processes and clinical outcomes

To understand how the reduction in fractional occupancy of the Salience-somatomotor substate was related to changes in psychological processes and clinical outcomes (Figure S5), we correlated changes in the Salience-somatomotor substate with significant group by time changes in questionnaire scores. We found associations with increased decentering, interoceptive awareness (listening to the body for insight) and reduced depressive symptoms (at both pre-to post-treatment and pre-treatment to 3 months follow up) when looking at the whole sample. Going a step further and exploring associations specific to the MBCT+TAU group and not the TAU control group, only the association between reduced fractional occupancy of the Salience-somatomotor substate and reduced depressive symptoms in the MBCT+TAU group at post-treatment and at 3 months follow up was specific to the MBCT+TAU group (See supplementary Table S7). For associations of change in lifetimes of the global state during rumination to psychological processes and clinical outcomes see the supplemental results.

To explore differential effects in the MBCT+TAU and TAU groups, we tested this in interaction models. For depressive symptoms, the association with fractional occupancy of the Salience-somatomotor substrate was significantly different between MBCT+TAU and TAU at post-treatment (Beta=0.41, t=1.9, p=0.03) and at 3 months (Beta = 0.55; t = 2.5; p = 0.017). Post hoc, this was explained by a reduction in fractional occupancy of the Salience-somatomotor substrate following MBCT+TAU being associated with improved clinical outcomes, while in the TAU group this was not found (Figure 4). For decentering and interoceptive awareness this interaction term was not significant.

**Figure 4:**
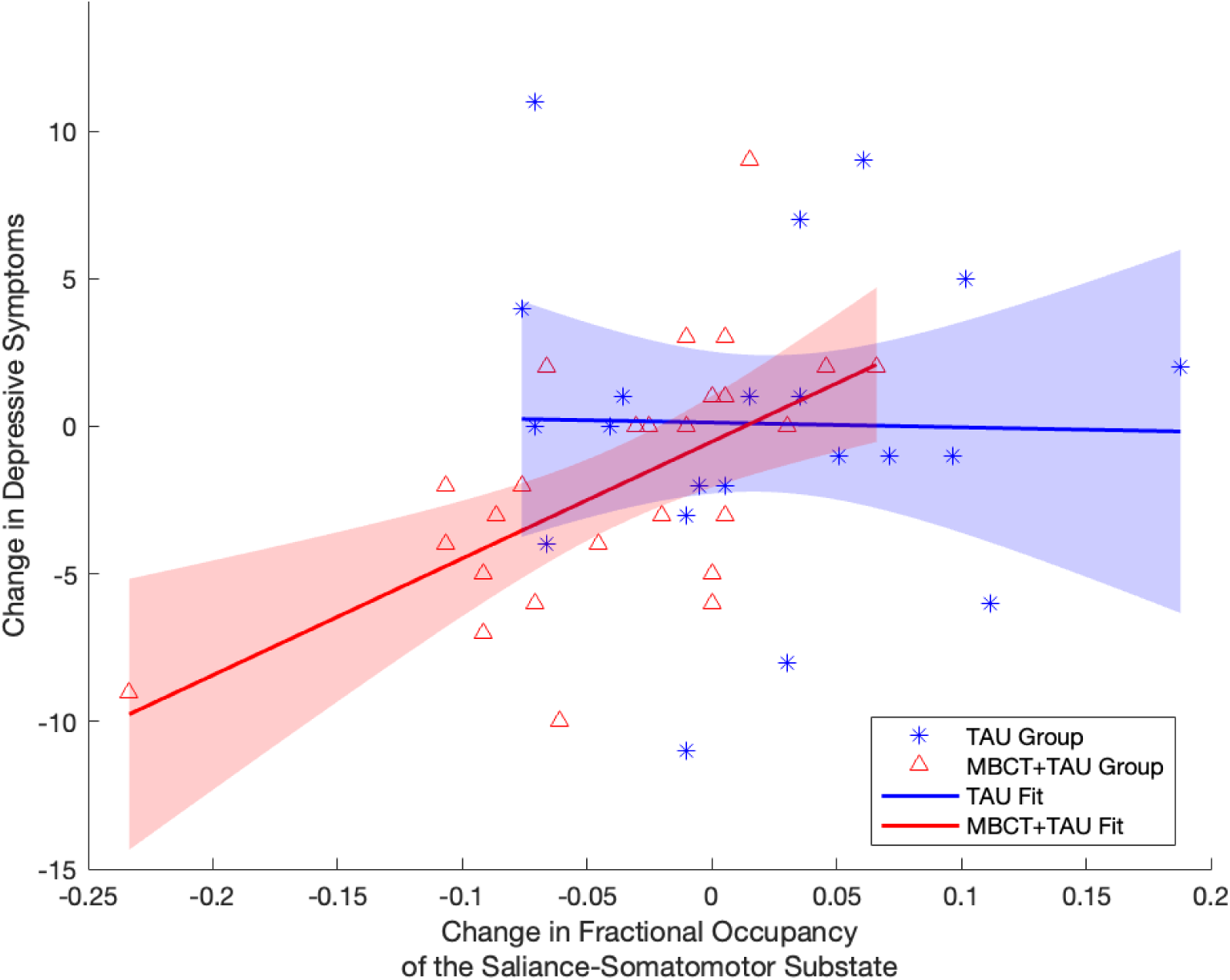
Changes in the Fractional Occupancy of the Salience-somatomotor substate post-treatment are associated with changes in depressive symptoms at 3 months of follow-up. Change in the Fractional Occupancy of the Salience-somatomotor substate (post-treatment - pre-treatment) in the MBCT+TAU group is significantly associated with changes in the depressive symptoms (3 months - pre-treatment), while this was not the case for the TAU group. The figure shows the linear fit of both MBCT+TAU and TAU groups with the 95% confidence intervals. Only the association between reduced fractional occupancy of the Salience-somatomotor substate and reduced depressive symptoms in the MBCT+TAU group post treatment and at 3 months follow up was specific to the MBCT+TAU group.

Of importance, the association between Fractional Occupancy of the Salience-Somatomotor substrate and severity of depressive symptoms was not present at baseline; neither in the whole group (Beta=0.08; t=0.54, p=0.59), nor for the two groups separately (MBCT+TAU allocated: (Beta=0.05; t=0.23, p=0.82), TAU allocated: (Beta=0.11; t=0.44, p=0.59)), suggesting that the observed association is specific to the treatment effect, and not an artefact of change in severity of clinical symptoms.

### Relating change in lifetimes during rumination to psychological processes and clinical outcomes

The trend towards increased duration (Lifetimes duration) of the global state following MBCT+TAU treatment (significant before correction for multiple comparisons; Figure 2B) was associated with increases in decentering, mindfulness, and improved clinical outcomes (i.e. reduced depressive symptoms) at post-treatment and with improved clinical outcomes at 3 months follow up across the whole sample (see supplementary files). These associations were not present at baseline either in the whole group or when split into the two groups. However, when analysing whether effects were specific to the MBCT+TAU group, only increased duration of the global state and clinical outcome at three months follow up was specific to the MBCT+TAU group.

## Discussion

In a randomised controlled trial comparing the effect of MBCT+TAU versus TAU-only in individuals with recurrent depression, we examined the dynamic functional connectivity of brain networks during a ruminative state. We found that MBCT+TAU altered the probability of occurrence (Fractional Occupancy) of a dynamic functional substate comprising areas of the salience network, reward network, and somatosensory network, during a ruminative mind state, and these dynamic changes in turn were associated with improved clinical outcomes after treatment and at three months follow up. While we did also find a trend in MBCT+TAU altering the duration (Lifetimes) of substates, this pattern was not robust as it did not survive correction for multiple comparisons.

The findings are consistent with a growing body of literature implicating the salience network in depressive symptomatology and treatment response (16,17,32,33). Changes in areas of the salience network such as the insula have been linked to prediction of treatment response (34) across several forms of psychotherapy for depression, as well as treatment response to mindfulness-based interventions across a wide group of populations (35,36). In addition, the salience network has been suggested to play a key role in valence, affective biases, and persistence of depressive rumination (2). We also found areas of the somatosensory and motor networks (i.e rolandic operculum, supramarginal gyrus, frontal inferior operculum, temporal superior gyrus, central gyrus, pallidum), and reward network (e.g. putamen in the basal ganglia) which together have been implicated in reward and motivational circuits in depression, emotional processing and severity of depressive symptoms and recurrence risk (37,38); (39))

In contrast to previous static analysis (18), this dynamic approach enabled us to show an association of decreases in the fractional occupancy of this salience-somatomotor network with improved clinical status i.e. reduced depressive symptoms after treatment and at 3 months follow up. As with the previous static connectivity analysis (18), we did not find dynamic connectivity changes in the Default mode network connectivity. MBCT+TAU targets the ability to recognize, decenter, and disengage from ruminative thoughts, thus reducing the risk of spiralling downward into a depressive mood and the potential onset of relapse, rather than changing ruminative thought frequency or content (11). It has been proposed that the default mode network and the salience mode network may play different roles in depressive rumination, with the salience network being more related to persistence, “stickiness,” and inability to disengage from the ruminative state persistence, and the DMN being more related to the self-referential thought content (2). Hence, it may not so much be the prevalence of negative self-related thought content that changes during a ruminative state, but rather the persistence and stickiness of the ruminative state. We speculate that the reduced fractional occupancy change in areas of the salience-somatomotor substate may play a role in the persistence of depressive ruminative states and as such might contribute to improved clinical outcomes.

The study had a number of limitations. First, our choice of TAU as the control group is both a strength (generalizability, external validity) and a limitation (lack of specificity in the control condition). However, given the absence of a specific active control group, we cannot know whether the treatment effects are specific to MBCT+TAU treatment or whether other treatments may yield similar dynamic effects. Future research could address this by comparing MBCT with equally effective treatments and examine the extent to which the mindfulness meditation components of MBCT drive the dynamic connectivity change by using either a dismantling design or active attention control. Second, we used highly experienced MBCT teachers fulfilling internationally recognized good practices guidelines for teachers, trainers, and supervisors of mindfulness courses (26). However, we did not directly measure their adherence to the treatment protocol and teaching competency (e.g., Mindfulness-Based Interventions: Teaching Assessment Criteria) during the programs (40). Third, out of ethical concerns, participants could choose to opt out of the rumination condition and participate in the rest of the study. As those choosing not to participate in the rumination condition had higher symptoms at baseline, but did not differ on other measures, the changes in the dynamic connectivity in the rumination state may mainly refer to those with fewer residual symptoms and milder symptoms. Finally, while the LEiDA analytical methodology has been demonstrated as a useful clinical tool in describing and distinguishing brain dynamics in various populations (22,24,38,41), it cannot delineate the hierarchical role of the implicated substates. Hence, future work could extend this work by embedding novel approaches in understanding hierarchical and gradient-like spatiotemporal organisation in health and diseases (42)(43,44).

In conclusion, MBCT+TAU compared with TAU led to decreased fractional occupancy in a substate consisting of areas of the salience, somatomotor and reward network during a ruminative state, and this dynamic change was associated with reduced depressive symptoms post treatment and at 3 months follow up, and as such might play a mechanistic role in improved treatment outcomes.

## Data and code availability

The data from this study will be made available from the corresponding author upon reasonable request. FSL software is available by the authors at https://fsl.fmrib.ox.ac.uk/fsl/fslwiki/FslInstallation and LEiDA scripts at GitHub (https://github.com/PSYMARKER/leida-matlab).

## Data Availability

The data from this study will be made available from the corresponding author upon reasonable
request. FSL software is available by the authors at https://fsl.fmrib.ox.ac.uk/fsl/fslwiki/FslInstallation and LEiDA scripts at GitHub (https://github.com/PSYMARKER/leida-matlab).

https://fsl.fmrib.ox.ac.uk/fsl/fslwiki/FslInstall

## Acknowledgments

This research was funded by a Carlsberg Foundation Internalization Fellowship (CF21_0645) to AMvdV. JV is supported by EU H2020 FET Proactive project Neurotwin grant agreement no. 101017716. GC was supported by a Research Fellowship from the Netherlands Organization for Health Research and Development (ZonMw, grant number 636320016) and a Brain and Behavior Research Foundation (BBRF) Young Investigator Grant (grant number 29875). The views expressed in this publication are those of the authors and do not necessarily reflect those of the funders. AMvdV developed the original design with advice from WK, and JV, MK, HR, GC advised on the dynamic analysis strategy and methods. AMvdV was responsible for the management of the study and collection of the data. AMvdV, JV, HR, GC and MK created the analysis strategy. JV analyzed the MRI data with the LEiDA methodology, and AMvdV analyzed the correlations with the self-report data and clinical data. All authors contributed to the interpretations of the data. AMvdV and JV wrote the initial draft of the methods, results, and AMvdV wrote the initial draft of the introduction and discussion. All authors contributed to and approved the final manuscript.

## References

1. World Health Organization (2022): Depression and Other Common Mental Disorders: Global Health Estimates.

2. Borders A (2020): Rumination and Related Constructs: Causes, Consequences, and Treatment of Thinking Too Much. Academic Press.

3. Papageorgiou C, Wells A (2004): Depressive Rumination: Nature, Theory and Treatment. John Wiley & Sons.

4. Langenecker SA, Westlund Schreiner M, Bessette KL, Roberts H, Thomas L, Dillahunt A, et al. (2024): Rumination-Focused Cognitive Behavioral Therapy Reduces Rumination and Targeted Cross-network Connectivity in Youth With a History of Depression: Replication in a Preregistered Randomized Clinical Trial. Biol Psychiatry Glob Open Sci 4: 1–10.

5. Fang Y (2019): Depressive Disorders: Mechanisms, Measurement and Management. Springer Nature.

6. Kuyken W, Warren FC, Taylor RS, Whalley B, Crane C, Bondolfi G, et al. (2016): Efficacy of Mindfulness-Based Cognitive Therapy in Prevention of Depressive Relapse: An Individual Patient Data Meta-analysis From Randomized Trials. JAMA Psychiatry 73: 565–574.

7. Segal ZV, Dimidjian S, Beck A, Boggs JM, Vanderkruik R, Metcalf CA, et al. (2020): Outcomes of Online Mindfulness-Based Cognitive Therapy for Patients With Residual Depressive Symptoms: A Randomized Clinical Trial. JAMA Psychiatry 77: 563–573.

8. Farb N, Anderson A, Ravindran A, Hawley L, Irving J, Mancuso E, et al. (2018): Prevention of relapse/recurrence in major depressive disorder with either mindfulness-based cognitive therapy or cognitive therapy. J Consult Clin Psychol 86: 200–204.

9. Montero-Marin J, van der Velden AM, Kuyken W (2024): Mindfulness-Based Cognitive Therapy’s Untapped Potential. JAMA Psychiatry 81: 1059–1060.

10. Segal Z, Williams M, Teasdale J (2018): Mindfulness-Based Cognitive Therapy for Depression, Second Edition. Guilford Publications.

11. Segal ZV, Williams JMG, Teasdale JD (2012): Mindfulness-Based Cognitive Therapy for Depression. Guilford Press.

12. Young KS, van der Velden AM, Craske MG, Pallesen KJ, Fjorback L, Roepstorff A, Parsons CE (2018): The impact of mindfulness-based interventions on brain activity: A systematic review of functional magnetic resonance imaging studies. Neurosci Biobehav Rev 84: 424–433.

13. Tang Y-Y, Tang R (2020): The Neuroscience of Meditation: Understanding Individual Differences. Academic Press.

14. Tang Y-Y, Hölzel BK, Posner MI (2015): The neuroscience of mindfulness meditation. Nat Rev Neurosci 16: 213–225.

15. Vignaud P, Donde C, Sadki T, Poulet E, Brunelin J (2018): Neural effects of mindfulness-based interventions on patients with major depressive disorder: A systematic review. Neurosci Biobehav Rev 88. 10.1016/j.neubiorev.2018.03.004

16. Godlewska BR, Browning M, Norbury R, Igoumenou A, Cowen PJ, Harmer CJ (2018): Predicting Treatment Response in Depression: The Role of Anterior Cingulate Cortex. Int J Neuropsychopharmacol 21: 988–996.

17. Marwood L, Wise T, Perkins AM, Cleare AJ (2018): Meta-analyses of the neural mechanisms and predictors of response to psychotherapy in depression and anxiety. Neurosci Biobehav Rev 95: 61–72.

18. van der Velden AM, Scholl J, Elmholdt E-M, Fjorback LO, Harmer CJ, Lazar SW, et al. (2023): Mindfulness Training Changes Brain Dynamics During Depressive Rumination: A Randomized Controlled Trial. Biol Psychiatry 93: 233–242.

19. Preti MG, Bolton TA, Van De Ville D (2017): The dynamic functional connectome: State-of-the-art and perspectives. Neuroimage 160: 41–54.

20. Cabral J, Vidaurre D, Marques P, Magalhães R, Silva Moreira P, Miguel Soares J, et al. (2017): Cognitive performance in healthy older adults relates to spontaneous switching between states of functional connectivity during rest. Sci Rep 7: 5135.

21. Lord L-D, Expert P, Atasoy S, Roseman L, Rapuano K, Lambiotte R, et al. (2019): Dynamical exploration of the repertoire of brain networks at rest is modulated by psilocybin. Neuroimage 199: 127–142.

22. Figueroa CA, Cabral J, Mocking RJT, Rapuano KM, van Hartevelt TJ, Deco G, et al. (2019): Altered ability to access a clinically relevant control network in patients remitted from major depressive disorder. Hum Brain Mapp 40: 2771–2786.

23. Vohryzek J, Deco G, Cessac B, Kringelbach ML, Cabral J (2020): Ghost Attractors in Spontaneous Brain Activity: Recurrent Excursions Into Functionally-Relevant BOLD Phase-Locking States. Front Syst Neurosci 14: 20.

24. Alonso Martínez S, Deco G, Ter Horst GJ, Cabral J (2020): The Dynamics of Functional Brain Networks Associated With Depressive Symptoms in a Nonclinical Sample. Front Neural Circuits 14: 570583.

25. Vohryzek J, Cabral J, Lord L-D, Fernandes HM, Roseman L, Nutt DJ, et al. (2022, July 4): Brain dynamics predictive of response to psilocybin for treatment-resistant depression. bioRxiv. p 2022.06.30.497950.

26. Crane RS, Kuyken W (2019): The Mindfulness-Based Interventions: Teaching Assessment Criteria (MBI:TAC): reflections on implementation and development. Curr Opin Psychol 28: 6–10.

27. Rush AJ, Trivedi MH, Ibrahim HM, Carmody TJ, Arnow B, Klein DN, et al. (2003): The 16-Item Quick Inventory of Depressive Symptomatology (QIDS), clinician rating (QIDS-C), and self-report (QIDS-SR): a psychometric evaluation in patients with chronic major depression. Biological psychiatry 54. 10.1016/s0006-3223(02)01866-8

28. Mehling WE, Price C, Daubenmier JJ, Acree M, Bartmess E, Stewart A (2012): The Multidimensional Assessment of Interoceptive Awareness (MAIA). PloS one 7. 10.1371/journal.pone.0048230

29. Fresco DM, Moore MT, van Dulmen MH, Segal ZV, Ma SH, Teasdale JD, Williams JM (2007): Initial psychometric properties of the experiences questionnaire: validation of a self-report measure of decentering. Behavior therapy 38. 10.1016/j.beth.2006.08.003

30. Tran US, Glück TM, Nader IW (2013): Investigating the Five Facet Mindfulness Questionnaire (FFMQ): construction of a short form and evidence of a two-factor higher order structure of mindfulness. J Clin Psychol 69: 951–965.

31. Thomas Yeo BT, Krienen FM, Sepulcre J, Sabuncu MR, Lashkari D, Hollinshead M, et al. (2011): The organization of the human cerebral cortex estimated by intrinsic functional connectivity. J Neurophysiol. 10.1152/jn.00338.2011

32. Fox KCR, Nijeboer S, Dixon ML, Floman JL, Ellamil M, Rumak SP, et al. (2014): Is meditation associated with altered brain structure? A systematic review and meta-analysis of morphometric neuroimaging in meditation practitioners. Neurosci Biobehav Rev 43: 48–73.

33. Downar J, Blumberger DM, Daskalakis ZJ (2016): The Neural Crossroads of Psychiatric Illness: An Emerging Target for Brain Stimulation. Trends Cogn Sci 20: 107–120.

34. Lynch CJ, Elbau IG, Ng T, Ayaz A, Zhu S, Wolk D, et al. (2024): Frontostriatal salience network expansion in individuals in depression. Nature 633: 624–633.

35. Guu S-F, Chao Y-P, Huang F-Y, Cheng Y-T, Ng H-YH, Hsu C-F, et al. (2023): Interoceptive awareness: MBSR training alters information processing of salience network. Front Behav Neurosci 17: 1008086.

36. Giommi F, Bauer PR, Berkovich-Ohana A, Barendregt H, Brown KW, Gallagher S, et al. (2023): The (In)flexible self: Psychopathology, mindfulness, and neuroscience. Int J Clin Health Psychol 23: 100381.

37. Knowland D, Lilascharoen V, Pacia CP, Shin S, Wang EH, Lim BK (2017): Distinct Ventral Pallidal Neural Populations Mediate Separate Symptoms of Depression. Cell 170. 10.1016/j.cell.2017.06.015

38. Alonso S, Tyborowska A, Ikani N, Mocking RJT, Figueroa CA, Schene AH, et al. (2023): Depression recurrence is accompanied by longer periods in default mode and more frequent attentional and reward processing dynamic brain-states during resting-state activity. Hum Brain Mapp 44: 5770–5783.

39. Rolls ET (2018): The Brain, Emotion, and Depression. Oxford University Press.

40. Crane RS, Eames C, Kuyken W, Hastings RP, Williams JMG, Bartley T, et al. (2013): Development and validation of the mindfulness-based interventions - teaching assessment criteria (MBI:TAC). Assessment 20: 681–688.

41. Martinez SA, Tyborowska A, Ikani N, Mocking RJ, Figueroa CA, Schene AH, et al. (2022, September 4): Segregation of dynamic resting-state reward, default mode and attentional networks after remitted patients transition into a recurrent depressive episode. medRxiv. p 2022.09.02.22279550.

42. Vohryzek J, Cabral J, Timmermann C, Atasoy S, Roseman L, Nutt DJ, et al. (2023, August 21): Harmonic decomposition of spacetime (HADES) framework characterises the spacetime hierarchy of the DMT brain state. bioRxiv. p 2023.08.20.554019.

43. Turbulent-like Dynamics in the Human Brain (2020): Cell Rep 33: 108471.

44. Kringelbach ML, Perl YS, Tagliazucchi E, Deco G (2023): Toward naturalistic neuroscience: Mechanisms underlying the flattening of brain hierarchy in movie-watching compared to rest and task. Science Advances. 10.1126/sciadv.ade6049

