## Supplementary material for "Dynamical repertoire of brain networks in mindfulness cognitive behavioural therapy during rumination: A randomized controlled trial": Dynamical repertoire of brain networks in mindfulness cognitive behavioural therapy during rumination_ A randomized controlled trial SI.pdf

### Extended Methods and Materials

#### MRI acquisition

Functional and structural images of the brain were acquired on a 3 Tesla Siemens Magneton Skyra 3T scanner (Siemens, Erlangen, Germany, software version Scout) using a 32-channel head coil, and using conventional acquisition parameters as specified below.

**Structural MRI:** A structural three-dimensional T1-weighted (3D-T1) scan was acquired with the following parameters: 176 slices covering the whole brain, TE (echo time)/TR(repetition time) = 3.8/2300 ms, inversion time = 31260 ms, flip angle = 8°, Field of View (FOV) = 256 mm, spatial resolution 1 × 1 × 1 mm3, Generalised Autocalibrating Partially Parallel Acquisitions (Grappa) = 2, and phase-encoding direction = AP.

**Functional MRI:** The duration of the rumination state was five minutes with 203 volumes of 2D gradient-echo EPI fMRI data were acquired with the following parameters: 52 ascending axial slices covering the whole brain, 3.8 × 3.8 × 3.8 mm3, FOV 192, Grappa = 2, Multiband = 2, TE/TR = 30/1480 ms, flip angle = 65°, and phase-encoding direction = AP.

#### FMRI preprocessing

We used FSL tools (https://fsl.fmrib.ox.ac.uk/fsl/docs/#/) for preprocessing. Preprocessing steps followed standard procedures and included: skull-stripping (BET tool), registering the functional to the structural image (FLIRT tool with default settings for Boundary-Based registration), registering the structural image to standard space (FNIRT tool with default settings for 12 degrees of freedom and warp-resolution of 10mm), motion correction (MCFLIRT tool) and spatial smoothing of the data with 5mm kernel. We used an independent component analysis (ICA)-based strategy for Automatic Removal of Motion Artefacts (ICA-AROMA). For further denoising, the first five eigenvariates of time courses extracted from white matter and cerebrospinal fluid masks (segmentation was done using FAST tool) were removed (using fsl_glm). Finally, data was high-pass filtered (100 seconds cut-off).

#### Leading Eigenvector Decomposition Analysis (LEIDA)

In this study, we applied Leading Eigenvector Decomposition Analysis (LEiDA) in order to elucidate dynamic changes in large-scale brain networks in patients undergoing MBCT treatment in ruminative brain states, and to describe the properties of the spatial substates, namely Fractional Occupancy (the percentage of frames assigned to a state for the whole of the rumination scan) and Life Times (the mean duration of temporally continuous runs of state occupancy). Figure 1 illustrates the experimental design consisted of comparing a group of patients undergoing MBCT in ruminative states compared to controls before and after treatment (Figure 1A) and the various steps involved in the analysis (Figure 1B). Across participants with both pre and post scans in the ruminative state, we transformed the parcellated fMRI signal into an analytical signal which allowed us to compute the phase relationship between each pair of regions at every recorded time point of the pre and post conditions. We subsequently computed the leading eigenvectors of each phase-locking pattern (a pattern consisting of regions with similar phase) at every recorded timepoint. Then, applying an unsupervised clustering algorithm (a machine learning tool to find structure in the data), we defined the number of clusters based on i) optimal clustering performance as determined by a silhouette score (a control measure quantifying the cohesion of the clusters) and ii) the lowest clustering solution that would result in Bonferroni corrected statistical significance for the Fractional Occupancy measure. To thoroughly investigate different partitions, we gradually varied k from 2 to 20. This approach allowed us to analyse substates that exhibited significant differences in brain activity between the MBCT intervention and control groups. The number of clusters, k, in the k-means clustering algorithm determines the range of phase-locking (PL) substates in the repertoire. By increasing k, more intricate, infrequent, and asymmetric networks are revealed. Lastly we computed the dynamic measures (fractional occupancy and life times) for each phase-locking substate, comparing change from pre to post treatment in the MBCT and TAU groups ( as described in detail below).

**Phase-locking Dynamics**

We first obtained the analytical signal for each region n where n = 1… 90 for the length of its timeseries t. This was calculated by first band-passing the regional signal in a narrow-band of 0.01-0.1 Hz and applying the Hilbert transform. The analytical signal represents the signal in terms of its instantaneous amplitude and phase, ϴ. We use the phase information at every timepoint of the recording to obtain the instantaneous phase coherence matrix as follows: iPC(n,m,t) = cos(ϴ(n,t)-ϴ(m,t)). This simple relationship accounts for the level of phase alignment between individual brain regions. If cos(0) = 1 and the regions are fully phase aligned, if cos(π) = -1 and the regions are fully phase anti-aligned and if cos(π/2) = 0 the brain regions are phase orthogonal to each other.

**Leading Eigenvector Dynamics**

In order to obtain the leading eigenvector dynamics, we applied principal component decomposition (PCA) to each iPC and selected the eigenvector with the strongest contribution thus obtaining leading eigenvectors at every timepoint $V_1(t)$ of dimensions 1xN. It has been shown that the leading eigenvector represents at least 50% of the iPC at each timepoint.. Furthermore, each leading eigenvector represents a spatial distribution across the parcellation atlas and can be mapped onto the cortical rendering for visualisation.

**Clustering into cortical substates**

In order to represent the leading eigenvectors in terms of recurring substates of brain activity in time, we cluster them using the unsupervised learning k-means algorithm. To do so, we concatenated all the timeseries across subjects and conditions in order to achieve a common space where N was the number of dimensions and (TxSbjxCnd) was the number of datapoints. We used the cosine distance as a measure of similarity and ran the algorithm for a varying number of cluster solutions - from 2-20 clusters with 100 repetitions. Then, every leading eigenvector, representing specific timepoints, was assigned to a given cluster centroid. Lastly, each centroid in a given clustering solution was associated to a given Yeo et al. (2011) functional network by computing the spatial correlation between the functional networks and the centroids themselves.

**Dynamical Measures of Fractional Occupancy and Lifetimes**

Fractional Occupancy was calculated as a probability of occurrence of a given substate α (centroids) across the time recording. Formally, fractional occupancy was calculated as follows

$$\prod_{\alpha} = \frac{1}{T}\sum_{t=1}^{T} \chi[\bar{x}(t) \in R^{\alpha}]$$

with χ being the indicator function taken on 1 if the state is on at time t and 0 if this is not the case. Fractional occupancy was calculated for every subject and condition.

Furthermore, we computed Life Times that quantifies the amount of time a given subset α is consecutively active

$$LT_{\alpha} = \frac{1}{p_{\alpha}}\sum_{1}^{p_{\alpha}} C_{p_{\alpha}}$$

where $p_{\alpha}$ is the number of consecutive periods and $C_{p_{\alpha}}$ their duration).

**Table S1. Baseline characteristics per group**

| Category | MBCT+TAU (N = 50) | TAU (N = 30) |
|---|---|---|
| **Sociodemographic Characteristics** | **n = 48** | **n = 28** |
| **Age, Years** | 43.17 (14.22) | 45.25 (12.01) |
| **Sex, Female/Male** | 35/15 (70%) | 23/5 (82%) |
| **Educational Level** | | |
| Low (<2-year further education) | 15 (30%) | 3 (11%) |
| Medium (2–4-year further education) | 24 (48%) | 21 (75%) |
| High (>5-year further education) | 9 (18%) | 4 (14%) |
| **Marital Status** | | |
| Married/cohabiting | 43 (90%) | 21 (75%) |
| Single/Not cohabiting | 5 (10%) | 7 (25%) |
| **Occupational Status** | | |
| Employed | 24 (50%) | 14 (50%) |
| Unemployed/benefits | 10 (20%) | 4 (14%) |
| Student | 3 (6%) | 1 (4%) |
| Retired | 7 (15%) | 4 (14%) |
| Other | 9 (19%) | 5 (18%) |
| **Clinical Characteristics** | | |
| Symptomatic (QIDS > 5) | 43 (83%), n = 50 | 25 (76%), n = 28 |
| Antidepressant Usage | 43/7 (86%), n = 50 | 21/7 (75%), n = 28 |
| Childhood Trauma | 58.79 (6.22), n = 42 | 58.96 (6.33), n = 26 |
| Previous Episodes of Depression | 3.90 (1.44), n = 41 | 3.80 (1.36), n = 23 |
| **Outcomes** | **n = 48** | **n = 27** |
| QIDS | 9.23 (4.58) | 9.68 (5.10) |
| EQ | 31.43 (7.12) | 31.26 (7.06) |
| MAIA_AR | 17.22 (5.03) | 17.78 (4.99) |
| MAIA_BL | 6.25 (2.07) | 7.40 (3.25) |
| MAIA_TR | 8.89 (3.31) | 8.40 (3.77) |
| MAIA_NO | 12.79 (2.61) | 13.96 (3.38) |

| | | |
|---|---|---|
| MAIA_ND | 9.17 (2.64) | 9.01 (2.45) |
| MAIA_EA | 15.32 (3.51) | 16.57 (4.23) |
| FFMQ | 44.21 (8.88) | 45.33 (80.2) |
| RRS | 53.38 (9.80) | 57.51 (8.24) |

Values are presented as **n**, **n (%)**, and **mean (SD)**.

***Abbreviations****: AR, attention regulation; BL, body listening; EA, emotional awareness; EQ, Experience Questionnaire; FFMQ, Five Factor Mindfulness Questionnaire; MAIA, Multidimensional Assessment of Interoceptive Awareness; ND, not-distracting; NO, noticing; QIDS, Quick Inventory of Depressive Symptomatology.*

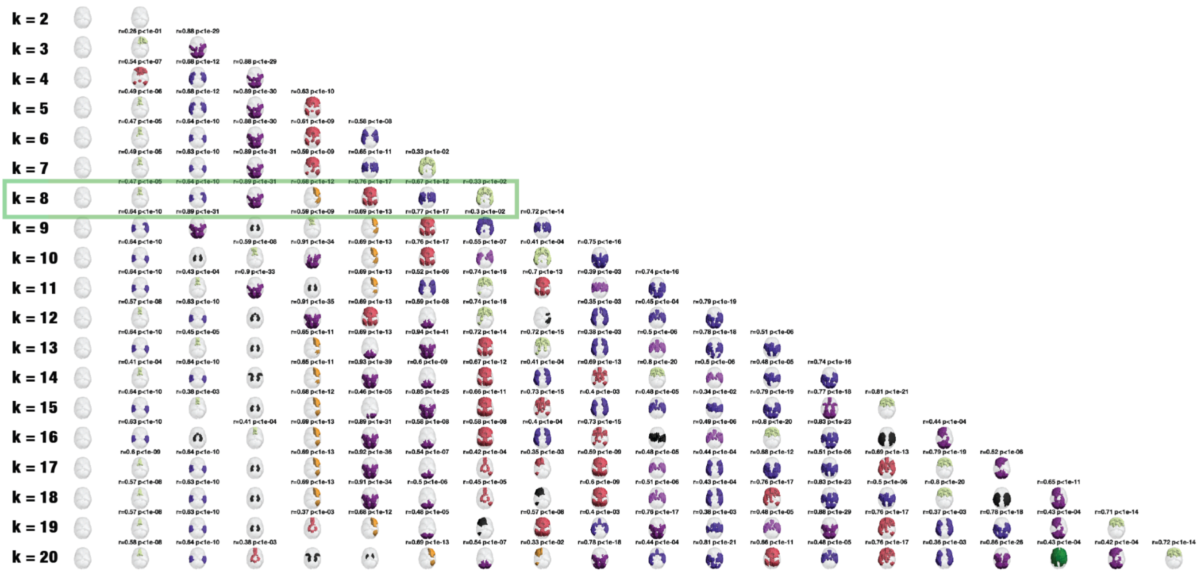


**Figure S2 - Pyramidal view of the clustering solutions.** Varying number of substates with increasing number solutions of the clustering algorithm. From k=2 to k=20. The substates are represented in cortical space for the AAL parcellation. The most significant resting-network has been associated (as defined by (Thomas Yeo et al. 2011) (31)) to each substate. We report the most significant resting-state network only when it survives the Bonferroni threshold $p < 0.05/K$ where K is the number of substates.

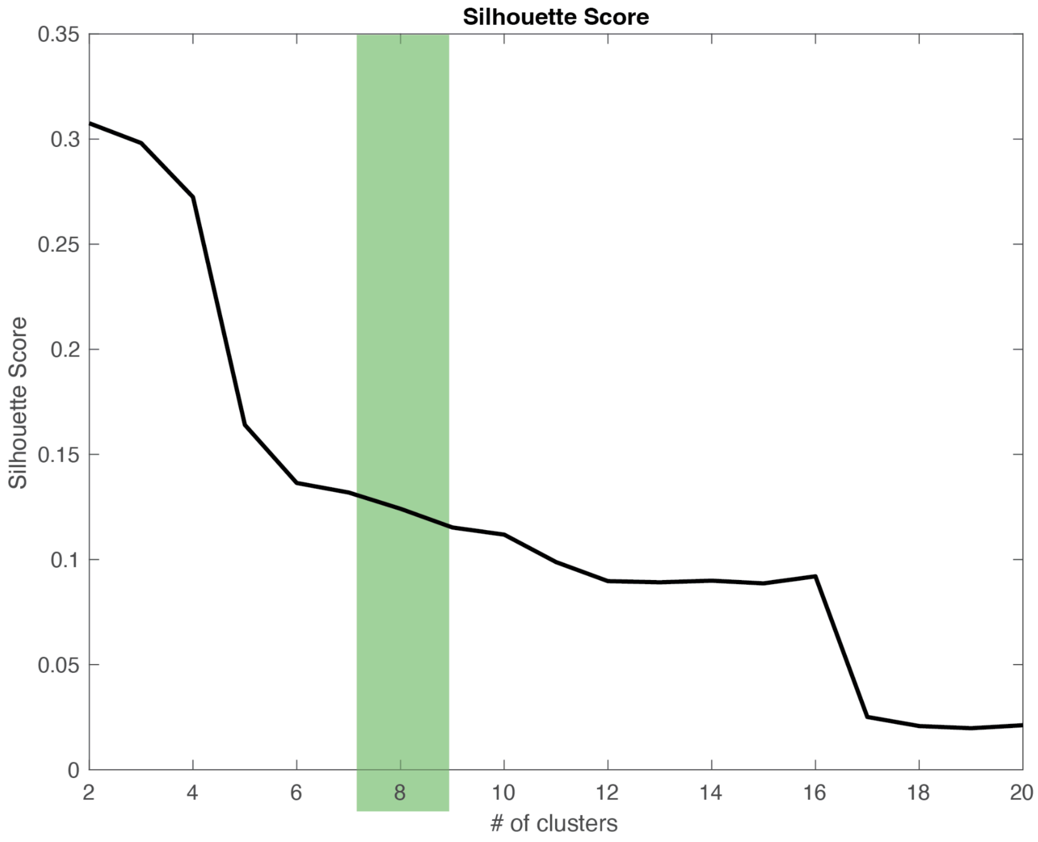


**Figure S3 - Cluster evaluation across the clustering solutions.** Silhouette scores evaluated for every clustering solution from k=2 to k=20.

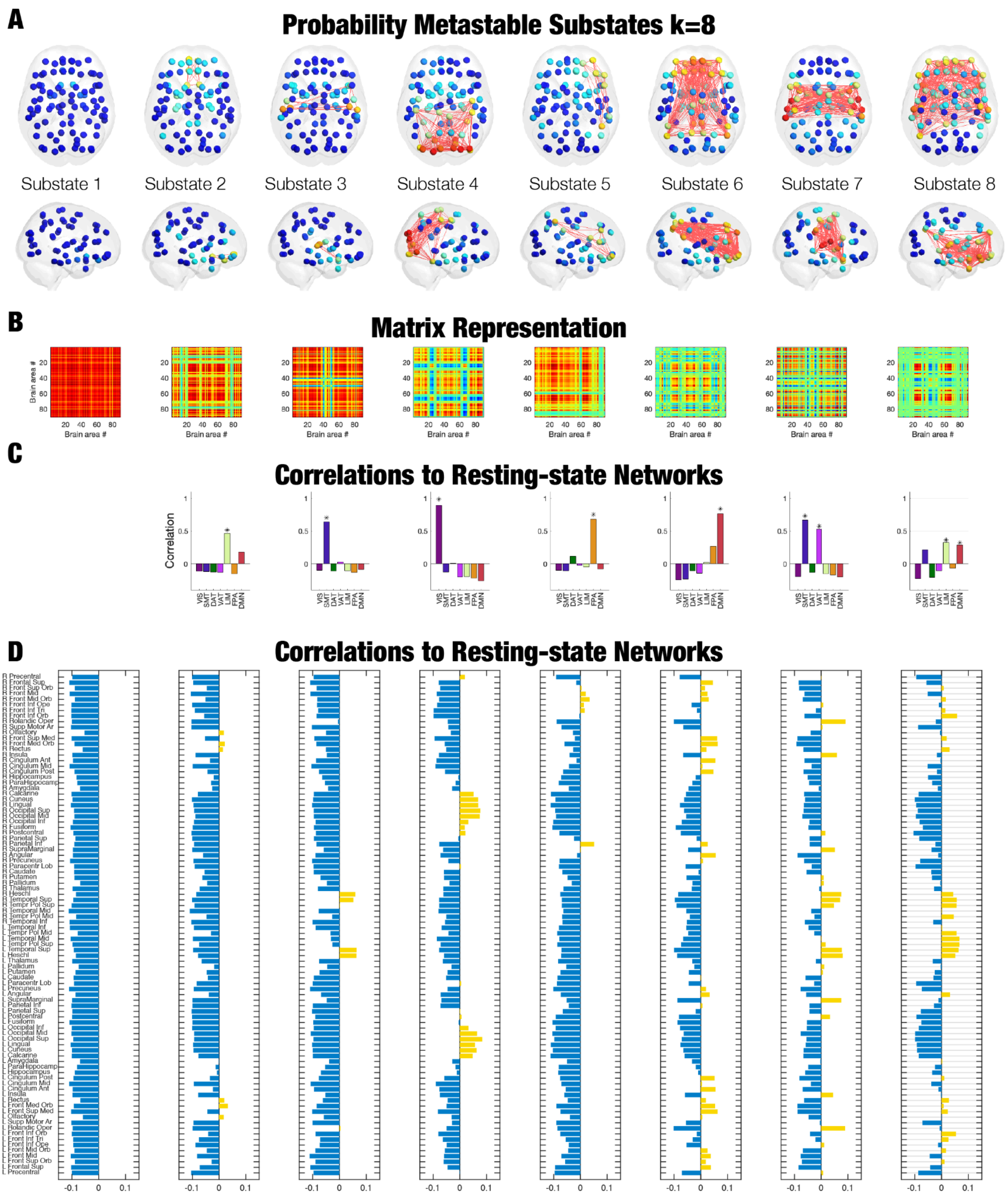


**Figure S4: Phase-locking solutions for 8 substates. A)** Cortical renders of the 8 substates. Here, we show the most important links connecting the given substates. **B)** Matrix representation of the PL substates. **C)** Association of the PL substates to the known 7 Resting-state networks as defined by Yeo et al. 2011 (31). **D)** Regional identification for all 8 PL substates.We chose the number of substates (8) based on the optimal clustering performance as determined by the silhouette score (Figure S3) as well as the lowest clustering solution that resulted in Bonferroni corrected statistical significance for the Fractional Occupancy measure..

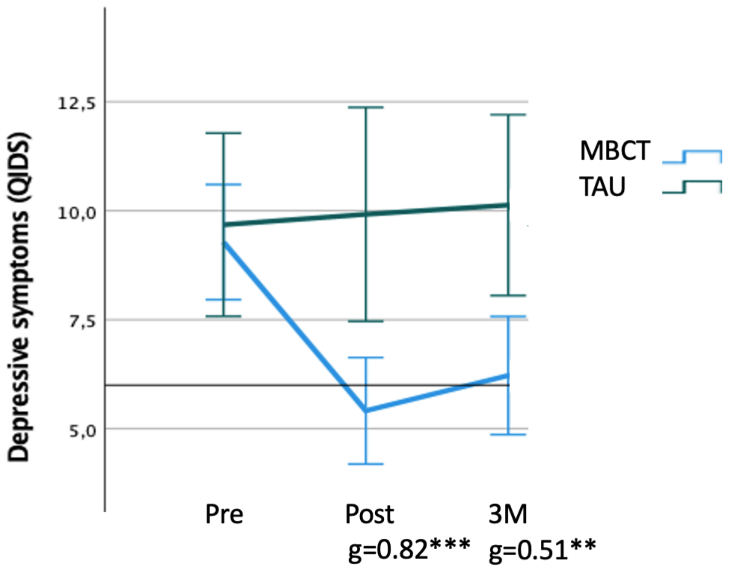


**Figure S5 - Clinical findings across time and group**

The MBCT+TAU group experienced a significant reduction in depressive symptoms (QIDS) at both post treatment (g=0.82, p>0.001), and at 3 months follow up (3M: g=0.51, p>0.0001), whereas no change was found in the TAU control group. Depressive symptoms measured by Quick Inventory of Depressive Symptoms (QIDS)<6 = below symptomatic threshold; QIDS 6-10 = Mild depressive symptoms; QIDS > 10 = moderate depressive symptoms.

**Table S7: Neural, psychological and clinical change score correlations across groups**

***WHOLE GROUP***

| . | | | | ***S1_LT*** | ***S5_LT*** | ***S7_FA*** |
|---|---|---|---|---|---|---|
| ***S1_LT*** | *Pearson Correlation* | | | *1* | *.101* | *.344* |
| | *Sig. (2-tailed)* | | | *.496* | *.017* | |
| | *N* | *48* | *48* | *48* | | |
| ***S5_LT*** | *Pearson Correlation* | | | *.101* | *1* | *.317* |
| | *Sig. (2-tailed)* | | *.496* | | *.028* | |
| | *N* | *48* | *48* | *48* | | |
| ***S7_FA*** | *Pearson Correlation* | | | *.344* | *.317* | *1* |
| | *Sig. (2-tailed)* | | *.017* | *.028* | | |
| | *N* | *48* | *48* | *48* | | |
| ***MAIA_NO*** | *Pearson Correlation* | | | *.016* | *-.168* | *-.042* |
| | *Sig. (2-tailed)* | | *.916* | *.276* | *.789* | |
| | *N* | *44* | *44* | *44* | | |
| ***MAIA_EA*** | *Pearson Correlation* | | | *-.184* | *-.200* | *-.021* |
| | *Sig. (2-tailed)* | | *.231* | *.194* | *.893* | |
| | *N* | *44* | *44* | *44* | | |
| ***MAIA_AR*** | *Pearson Correlation* | | | *-.237* | *-.026* | *-.283* |
| | *Sig. (2-tailed)* | | *.122* | *.865* | *.063* | |
| | *N* | *44* | *44* | *44* | | |
| ***MAIA_BL*** | *Pearson Correlation* | | | *-.237* | *-.298* | *-.365* |
| | *Sig. (2-tailed)* | | *.121* | *.050* | *.015* | |
| | *N* | *44* | *44* | *44* | | |
| ***QIDS_POST*** | *Pearson Correlation* | | | *.346* | *.132* | *.322* |
| | *Sig. (2-tailed)* | | *.018* | *.382* | *.029* | |
| | *N* | *46* | *46* | *46* | | |
| ***QIDS_3M*** | *Pearson Correlation* | | | *.409* | *.099* | *.378* |
| | *Sig. (2-tailed)* | | *.006* | *.521* | *.011* | |
| | *N* | *44* | *44* | *44* | | |
| ***EQ*** | *Pearson Correlation* | | | *-.379* | *-.320* | *-.441* |
| | *Sig. (2-tailed)* | | *.011* | *.034* | *.003* | |
| | *N* | *44* | *44* | *44* | | |
| ***FFMQ*** | *Pearson Correlation* | | | *-.378* | *-.164* | *-.260* |
| | *Sig. (2-tailed)* | | *.011* | *.286* | *.08* | |

***MBCT+TAU***

| | | | | | | |
|---|---|---|---|---|---|---|
| ***S1_LT*** | *Pearson Correlation* | | | *1* | *.101* | *.344* |
| | *Sig. (2-tailed)* | | | *.496* | *.017* | |
| | *N* | *27* | *27* | *27* | | |
| ***S5_LT*** | *Pearson Correlation* | | | *-.028* | *1* | |
| | *Sig. (2-tailed)* | | *.891* | | *.522* | |
| | *N* | *27* | *27* | *27* | | |
| ***S7_FA*** | *Pearson Correlation* | | | *.321* | *.129* | *1* |
| | *Sig. (2-tailed)* | | *.103* | *.522* | | |
| | *N* | *27* | *27* | *27* | | |
| ***MAIA_NO*** | *Pearson Correlation* | | | *.179* | *-.110* | *.301* |
| | *Sig. (2-tailed)* | | *.383* | *.593* | *.135* | |
| | *N* | *26* | *26* | *26* | | |
| ***MAIA_EA*** | *Pearson Correlation* | | | *-.121* | *.105* | *.270* |
| | *Sig. (2-tailed)* | | *.556* | *.608* | *.182* | |
| | *N* | *26* | *26* | *26* | | |
| ***MAIA_AR*** | *Pearson Correlation* | | | *-.158* | *.110* | *.014* |
| | *Sig. (2-tailed)* | | *.439* | *.593* | *.947* | |
| | *N* | *26* | *26* | *26* | | |
| ***MAIA_BL*** | *Pearson Correlation* | | | *-.165* | *-.260* | *-.090* |
| | *Sig. (2-tailed)* | | *.420* | *.199* | *.661* | |
| | *N* | *26* | *26* | *26* | | |
| ***QIDS_POST*** | *Pearson Correlation* | | | *.329* | *.064* | *.428* |
| | *Sig. (2-tailed)* | | *.094* | *.750* | *.026* | |
| | *N* | *27* | *27* | *27* | | |
| ***QIDS_3M*** | *Pearson Correlation* | | | *.415* | *-.040* | *.573* |
| | *Sig. (2-tailed)* | | *.031* | *.845* | *.002* | |
| | *N* | *27* | *27* | *27* | | |
| ***EQ*** | *Pearson Correlation* | | | *-.333* | *-.286* | *-.223* |
| | *Sig. (2-tailed)* | | *.097* | *.156* | *.274* | |
| | *N* | *26* | *26* | *26* | | |
| ***FFMQ*** | *Pearson Correlation* | | | *-.343* | *-.054* | *-.081* |
| | *Sig. (2-tailed)* | | *.086* | *.793* | *.693* | |
| | *N* | *26* | *26* | *26* | | |

| | | | | | | | |
|---|---|---|---|---|---|---|---|
| ***TAU*** | ***S1_LT*** | *Pearson Correlation* | | | *1* | *-.026* | *.103* |
| | | *Sig. (2-tailed)* | | | *.913* | *.656* | |
| | | *N* | *21* | *21* | *21* | | |
| | ***S5_LT*** | *Pearson Correlation* | | | *-.026* | *1* | *.276* |
| | | *Sig. (2-tailed)* | | *.913* | | *.226* | |
| | | *N* | *21* | *21* | *21* | | |
| | ***S7_FA*** | *Pearson Correlation* | | | *.103* | *.276* | *1* |
| | | *Sig. (2-tailed)* | | *.656* | *.226* | | |
| | | *N* | *21* | *21* | *21* | | |
| | ***MAIA_NO*** | *Pearson Correlation* | | | *.068* | *.066* | *-.251* |
| | | *Sig. (2-tailed)* | | *.788* | *.795* | *.315* | |
| | | *N* | *18* | *18* | *18* | | |
| | ***MAIA_EA*** | *Pearson Correlation* | | | *.108* | *-.237* | *.090* |
| | | *Sig. (2-tailed)* | | *.670* | *.344* | *.723* | |
| | | *N* | *18* | *18* | *18* | | |
| | ***MAIA_EA*** | *Pearson Correlation* | | | *.183* | *.341* | *-.431* |
| | | *Sig. (2-tailed)* | | *.467* | *.166* | *.074* | |
| | | *N* | *18* | *18* | *18* | | |
| | ***MAIA_BL*** | *Pearson Correlation* | | | *.171* | *-.053* | *-.396* |
| | | *Sig. (2-tailed)* | | *.498* | *.834* | *.104* | |
| | | *N* | *18* | *18* | *18* | | |
| | ***QIDS_POST*** | *Pearson Correlation* | | | *.061* | *-.017* | *-.109* |
| | | *Sig. (2-tailed)* | | *.803* | *.945* | *.657* | |
| | | *N* | *19* | *19* | *19* | | |
| | ***QIDS_3M*** | *Pearson Correlation* | | | *.125* | *.030* | *-.139* |
| | | *Sig. (2-tailed)* | | *.632* | *.910* | *.596* | |
| | | *N* | *17* | *17* | *17* | | |
| | ***EQ*** | *Pearson Correlation* | | | *-.037* | *-.075* | *-.394* |
| | | *Sig. (2-tailed)* | | *.884* | *.769* | *.105* | |
| | | *N* | *18* | *18* | *18* | | |
| | ***FFMQ*** | *Pearson Correlation* | | | *-.216* | *-.109* | *-.298* |
| | | *Sig. (2-tailed)* | | *.389* | *.668* | *.231* | |
| | | *N* | *18* | *18* | *18* | | |

***Table S7: Correlations between significant change scores in Lifetimes, Fractional Occupancy, psychological processes and clinical outcomes for the whole sample and MBCT+TAU and TAU separate.*** *Change scores (post treatment minus pre treatment or 3 months forllow minus pre treatment (only for QIDS_3M). Abbreviations: S1_LF Lifetimes substrate 1; S5_LT Lifetimes substrate 5; S7_FA Fractional Occupancy substrate 7 (I.e. Salience-somatomotor substrate); MAIA, Multidimensional Assessment of Interoceptive Awareness; ND, not-distracting; NO, noticing; AR, attention regulation; BL, body listening; EA, emotional awareness; QIDS, Quick Inventory of Depressive Symptomatology; EQ, Experience Questionnaire; FFMQ, Five Factor Mindfulness Questionnaire.*
